# A distributed, privacy-preserving platform for linkage of epidemiological data with pathogen genome sequences

**DOI:** 10.64898/2026.01.18.26344367

**Authors:** Jolene Langevin, Leo A. Featherstone, Francesca Di Giallonardo, Bethany A. Horsburgh, Ha Q. Vo, Andrew Lloyd, William Rawlinson, Rowena A. Bull, Anthony D. Kelleher, Lachlan J.M Coin, the H2Seq Data management working group

## Abstract

Efficient and secure integration of epidemiological data with pathogen genome sequence data is essential for identifying genetic clusters, monitoring emerging mutations and targeting public health responses. However, this information is often collected across different organisations: epidemiological data is collected by public health units while genome sequence data is collected by diagnostic laboratories. Linking these sources often requires manual or semi-manual approaches, leading to unnecessary delays in identifying emerging outbreaks. To address this, we developed a proof-of-concept privacy-preserving distributed platform, SecureEpiLink, for automatic linkage of pathogen genome sequences and notification data across public health and diagnostic laboratories. SecureEpiLink uses cryptographic hashing to establish linkage, without exposing personal identifying information. This ensures that the resulting linked data can be used to identify the emergence of genetic clusters without identification of individuals comprising each cluster, with the original identifiable data remaining stored at source units. We benchmarked SecureEpiLink against manual linkage and another linkage service using HIV and HCV datasets from New South Wales, Australia. SecureEpiLink performed similarly to manual linkage and outperformed previously used linkage algorithms, with all errors attributable to data entry errors in the underlying dataset. Lastly, we demonstrate how SecureEpiLink can be integrated with automated genomic epidemiology pipelines. SecureEpiLink is available from https://github.com/jolenefarrell/SecureEpiLink.

**Author summary:** Linkage of epidemiological data collected in public health units with pathogen genome sequence data generated by diagnostic labs in real-time is essential for targeted public health responses. However, linkage increases the risk of identifying individuals and in turn stigmatisation and even criminalisation (in the case of HIV). We have designed, implemented and tested a privacy preserving real-time system for linking pathogen genome sequence data with public health notification data across different jurisdictions.

## Introduction

Public health notification data provide critical information on the overall spread and demographic correlates of infectious disease [1]. Additionally, pathogen genome sequence data can be used to identify genetic clusters, their rate of growth, as well as the emergence of new variants of concern [2–4]. The linkage of genome sequence with public health notification data provides the most complete picture of the way in which a pathogen is spreading in the community, and which demographic and geographic groups are most at risk [5–7]. As such, centralised databases of pathogen sequence data and public health notification data such as AusTrakka [8] and GISAID [9] provide powerful resources for genomic epidemiology. For example, numerous human immunodeficiency virus (HIV) studies have used molecular epidemiology to identify key factors that link a genetic cluster, which can be further utilised to design effective public health interventions [3]. Notably, the US and Canada have implemented molecular epidemiology as part of their disease control programmes [10, 11]. However, linking notification and genome sequence data is particularly complex for stigma-associated pathogens or where transmission is criminalised, such as with HIV [12], hepatitis-C virus (HCV) [13] and Mpox [14]. Data linkage platforms must therefore preserve the security of personal identifying data for linkage to be both successful, viable and acceptable to the at-risk community.

Recent advances in distributed computing, cryptographic protocols and privacy-preserving record linkage provide a promising alternative to traditional centralised systems, which often require the transfer of sensitive and identifiable data into a single repository. Despite this, practical implementations of distributed or decentralised platforms are limited. This study provides a proof-of-concept for distributed and de-identified data linkage and sharing platform, SecureEpiLink, that enables linkage of public health notification data with pathogen genome sequence data. SecureEpiLink allows organisations to store and manage their own data while linkage is performed across the data network using cryptographic hashing instead of exposing patients’ personal identifying information.

We demonstrate use of SecureEpiLink using empirical HIV and HCV datasets from New South Wales (NSW), Australia. Pathogen sequence data for notifiable pathogens is generated by multiple public and private diagnostic labs across Australia [15]. Notification data, which contains information on the demography of the patient, including age, their postcode and ethnicity, is collected by public health units, and typically stored within the Department of Health in each state jurisdiction. There are linkage services such as Centre for Health Record Linkage (CHeReL) [16] in NSW, which link pathogen genome sequence with notification data using an established deterministic algorithm. However, linkage takes place at arbitrary intervals due to request processes and financial investment requirements, limiting the ability for real-time analysis. Here, we show that SecureEpiLink achieves comparable-or-higher linkage success to deterministic linkage run at CHeReL for empirical HIV and HCV datasets. We further demonstrate use of SecureEpiLink with a simulated dataset, showing that all linkage failures are attributable to errors and inconsistencies in underlying datasets. We also demonstrate how SecureEpiLink can be integrated with automated bioinformatics pipelines. Lastly, we discuss considerations for how distributed platforms such as SecureEpiLink can be further developed and deployed at scale.

## Methods

### Overview

Below, we describe SecureEpiLink’s linkage procedure, as well as how we compared its performance to CHeReL and manual linkage using empirical and simulated datasets. We then demonstrate how SecureEpiLink can integrate with bioinformatic and data-visualisation pipelines. We evaluate linkage success for HIV by using records from the NSW HIV resistance database from 2004-2023 and NSW Notifiable Conditions Information Management System (NCIMS) database, and for HCV by using data from the SToP-C [17] and HITS-p [18] studies of HCV among prisoners (ethics approval 2024/ETH00857). We then use another previously published NSW HIV genomic dataset [19] with randomised metadata to demonstrate how SecureEpiLink can integrate with reporting pipelines.

### Description of linkage protocol

We used a one-way cryptographic hash function to de-identify and link notification and genomic datasets. A linkage identifier was generated for each record by hashing concatenated identifiable fields and appending a randomly generated string, also known as a salt. Notification and genomic records were then matched on the generated linkage identifiers. We assumed that there was a single genomic-notification record pair, where successful linkage was defined as one genomic record matching one notification record uniquely. Failed linkage was defined as either undetermined, where a genomic or notification record matched multiple records in the corresponding dataset due to those records sharing the same linkage identifier (also referred to as a linkage collision), or unsuccessful, where one genomic or notification record matched no records in the corresponding dataset.

### Distributed data linkage platform

We developed a proof-of-concept platform, SecureEpiLink, to showcase the utility of the linkage protocol. SecureEpiLink was implemented using Node.js (v24) and the Express web application framework, with development carried out in an isolated test environment, separate from any production systems, allowing for modular development, experimentation and quick prototyping. Isolated environments allow for modular development and prototyping as all the components can be built and tested without affecting real data or systems. We designed API endpoints to facilitate data linkage using standard HTTP methods, including GET and POST, and simple authentication was handled through shared API keys included in all request headers. SecureEpiLink was containerised with Docker, a process that allows all code and external dependencies to be packaged together in an isolated lightweight environment. All relevant data was stored in a MySQL database, and SecureEpiLink was configured to produce structured JSON logs. We used 347 previously published HIV genomes [19] (GenBank accession numbers MW167298 – MW167644) for which we simulated notification times and key metadata categories, including personal-identifiers, to test the platform. Metadata (Supplementary Table 1) and the corresponding notification dataset (Table 7) was generated utilising a custom R script. The key notification and genomic metadata fields and value distributions (Supplementary Table 2 and 3) were guided by the annual NSW HIV Strategy 2021-2025 Annual Data Report 2024 [20]. To simulate real-world data collection and entry errors, we introduced errors into the name codes (the first two letters of an individual’s first name combined with the first two letters of the individual’s surname) of 17 out of the 347 records (5%) in the genomic dataset. These records were selected at random, and in each the two letters from either the first two characters of the first name, or first two characters of the surname, were reversed. A fixed random seed was used so the procedure is reproducible, with the simulation script made available in the project repository [21].

### Participating linkage fields analysis

To evaluate which identifiable fields provided the highest linkage success for both HIV and HCV datasets, we analysed multiple different combinations of fields as input to the anonymised linkage identifier. The identifiable fields included name code, date of birth, sex, postcode, sexual health clinic code (unique identifier from the attended sexual health clinic) and incarceration facility (prison) (Supplementary Table 4 and 5). These variables were selected based on available data sources for testing, where a smaller genomic dataset was linked with a larger conclusive notification dataset. Following the selection of candidate linkage fields, the approach was applied to a HIV genomic dataset, consisting of a random subset (n=276) of records selected from the NSW Resistance Database from 2011 to 2021. The corresponding HIV notification dataset (n=22,187) was extracted from the NCIMS database. The HCV genomic dataset (n=633) and corresponding notification dataset (n=2410) were derived from the Surveillance and Treatment of Prisoners with Hepatitis C (SToP-C) [17], and Hepatitis C Incidence and Transmission Study in prisons (HITS-p) [18] cohorts. The number of missing values for both the notification and genomic dataset for both pathogens are shown in Table 1, noting that missing counts could not be reported for HIV notification metadata records. Notification and genomic datasets for both pathogens were linked deterministically using a custom Python script, where linkage was repeated for a defined set of identifiable field combinations, or linkage IDs. The Python script performed a series of independent linkage iterations, one for each combination of anonymised sensitive linkage fields. An overall linkage summary for each iteration was generated in CSV format for interrogation of successful, undetermined or unsuccessful linkage results. This script is not publicly available online, or released under a permissive license, as it is being utilised within organisations with strict security requirements that restrict publishing code online. The node.js implementation of SecureEpiLink linkage protocol is equivalent and generates the same results as the Python script given the same input data.

**Table 1.** Missing value counts from HIV and HCV genomic and notification datasets.

|  | **HIV** |  | **HCV** |  |
|---|---|---|---|---|
| **Field** | Genomic<br>(n=276) | Notification<br>(n=22187) | Genomic<br>(n=633) | Notification<br>(n=2140) |
|---|---|---|---|---|
| Name code | 0 | - | 7 | 7 |
| D.O.B | 0 | - | 7 | 7 |
| Sex | 0 | - | 0 | 0 |
| Postcode | 0 | - | Not used | Not used |
| Prison | Not used | Not used | 3 | 0 |

### Data linkage performance across methods

To further validate the developed data linkage protocol and assess the accuracy and efficiency of linkage utilising real data, we compared the proposed data linkage method with a well-established linkage protocol run at the NSW Centre for Health Record Linkage (CHeReL). This linkage protocol uses a series of deterministic passes matching on fields including date of birth, sex, first two letters of first name and surname, and postcode where available. References to CHeReL through the manuscript refer to the deterministic linkage algorithm run at the organisation. The previously described HIV genomic (n=276) and notification (n=22,187) datasets were utilised for this validation, using name code and date of birth as the inputs to the anonymised linkage identifier, where successful linkage of genomic records to notification records of both methods were compared.

We did not clean datasets prior to analysis with the aim of incorporating realistic variability in the data. Therefore, factors such as missing fields, inconsistent formatting and data typing are expected to have a significant influence on the linkage success.

### Simulated dataset linkage analysis

To investigate the impact of the uniqueness of linkage fields on linkage efficacy, we generated larger randomly generated datasets. Datasets of various sizes, containing only name codes and dates of birth, were generated with a custom R script utilising the randomNames package (version 1.6-0.0) [22] to generate first and last names. Simulated names generated from the package contain apostrophes, hyphens and spaces, although do not contain any accents or diacritics.

To mimic realistic populations, each record consisted of a date of birth between 1950 and 2010, and a name code based on ethnicity definitions within the package. To assess the effect of cultural diversity two versions of each dataset were generated, with the first version sampling first and last names from a single ethnic group and the second sampling from multiple ethnic groups, as defined by the package’s names populations. The assigned sampling probabilities for the ethnically diverse datasets were approximated using the 2021 NSW Census [23] as a guide (Table 2). To account for probable data entry errors in real-world applications, the characters of 16.67% of the name codes in each dataset were reversed. For each of the conditions, defined by dataset size and ethnic category, the occurrence of duplicate name code and date of birth combinations throughout each dataset was calculated and averaged over 10 repeated runs (Supplementary Table 6).

**Table 2.** Ethnic sub-sampling ratios used, based on 2021 NSW census statistics, as defined by the randomNames R software package. White effectively refers to Anglo-Saxon names.

| **NSW 2021 Census** |  | **randomNames R software package** |  |
|---|---|---|---|
| Ethnicity definition | Occurrence percentage | Ethnicity definition as in software package | Sampling probability percentage |
| English | 29.8% | White (not Hispanic) | 75% |
| Australian | 28.6% |  |  |
| Irish | 9.1% |  |  |
| Scottish | 7.7% |  |  |
| Chinese | 7.2% | Asian or Pacific Islander | 7% |
| Other | 17.6% | Other (random selection of all available ethnicities) | 18% |

### Report generation pipeline

To illustrate the applicability of the data linkage platform in future genomic epidemiology and public health initiatives, we developed a minimal integrated bioinformatic analysis and reporting component for the HIV simulation dataset. RESTful Python APIs were developed to run simple SnakeMake bioinformatic analysis pipelines and manage dynamic report generation with Quarto [24], utilising the linked dataset generated by the linkage platform.

Within the HIV bioinformatic pipeline (Figure 1A) a pairwise distance matrix was constructed from HIV sequences aligned to the HXB2 protease and reverse-transcriptase reference sequence (GenBank Accession number K03455.1), a common reference. The pairwise distance matrix was then used to generate a network visualisation JSON output which defines nodes, edges and clusters using HIV-TRACE (version 0.9.2) [25, 26] parameters (Supplementary Table 3). Note that genetic clusters defined on genetic distance do not necessarily indicate transmission events and only represent genetically similar infections. We also developed a pathogen-specific cluster report template in HTML format using Quarto. The report templates required the network visualisation output from the analysis pipeline, and the linked sequence and notification metadata stored in the persistent MySQL database (Figure 1B). Quarto templates used Plotly [27] (version 6.1.2) and networkx [28] (version 3.3) for visualisation.

**Figure 1.**
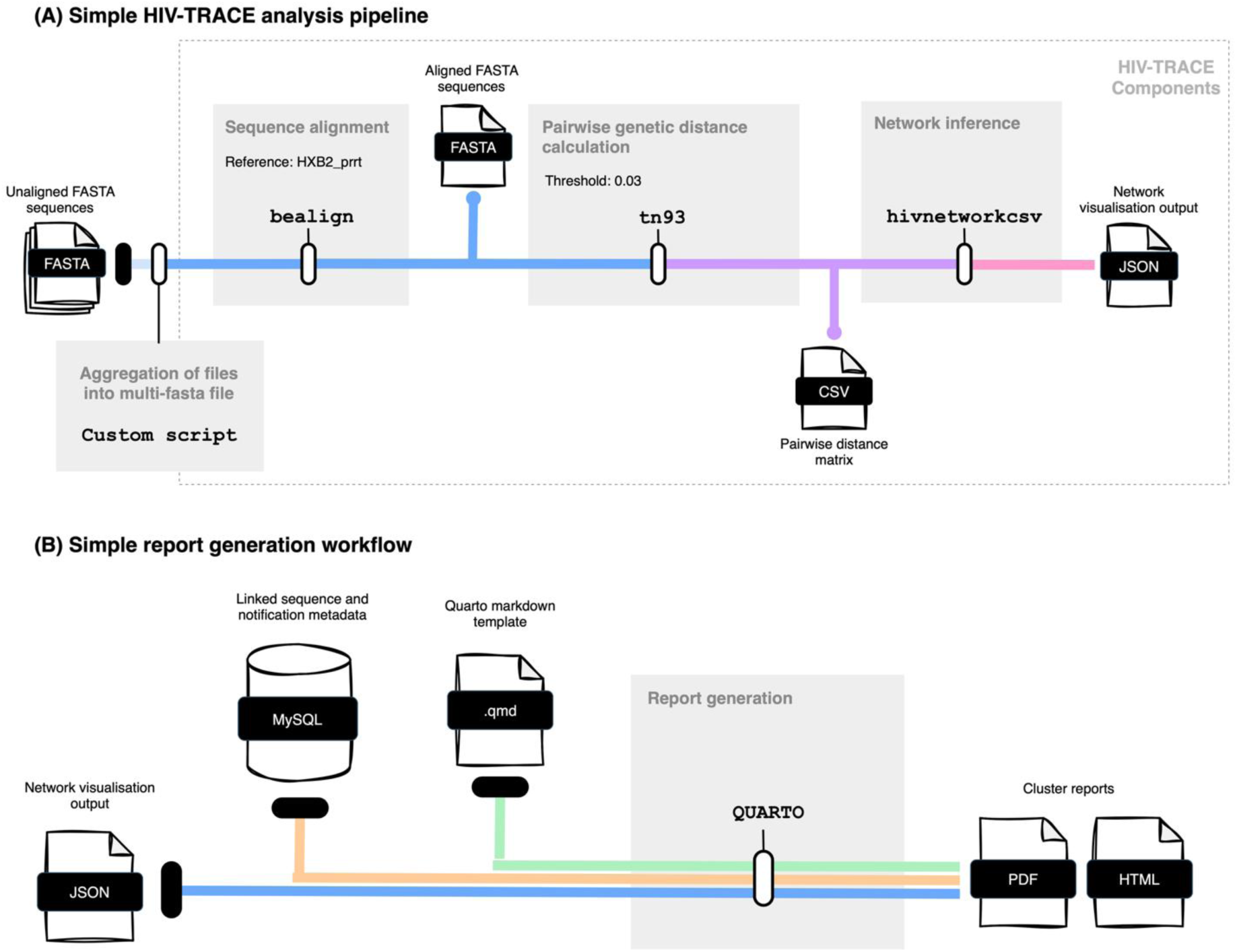
HIV Pipeline and reporting workflow. Schematic of the simple analysis workflow using HIV-TRACE (A) and reporting workflow using Quarto templates (B).

## Results

### Simulated dataset linkage analysis

To investigate the occurrence of duplicate name code and date of birth combinations, datasets of various sizes were generated to simulate real-world name codes and date of births over a defined period between 1/01/1950 and 1/01/2010. Increasing the dataset size by a factor of 10 resulted in a linear increase in the number of duplicated name code and date of birth combinations, illustrated as a percentage of total records (Table 3). The percentage of duplicates increases approximately 10-fold per 10-fold increase in records, and the raw count of duplicates rise approximately 100-fold per 10-fold increase in records.

**Table 3.** Duplicate name count using randomly generated first name, last name and date of birth combinations to simulate real-world data. Duplicate averages were calculated over 10 runs of randomly generated datasets of each size.

|  | **Caucasian-only names** |  |  | **Culturally diverse names** |  |  |
|---|---|---|---|---|---|---|
| Records | Duplicates (average) | Duplicates (percentage) | Standard deviation | Duplicates (average) | Duplicates (percentage | Standard deviation |
| 12,000 | 0.6 | 0.01% | 1.35 | 0 | 0.00% | 0.63 |
| 120,000 | 109.8 | 0.09% | 17.11 | 100 | 0.08% | 14.05 |
| 1,200,000 | 10,724 | 0.89% | 166.44 | 10,330 | 0.86% | 137.56 |
| 1.2 × 10<sup>6</sup> | 985,863 | 8.22% | 1,659.75 | 945,084 | 7.88% | 1,393.23 |

### Data linkage performance across methods

In a comparison of linkage methods linking records on name code and date of birth, SecureEpiLink achieved a higher linkage success result (Table 4), linking 241 out of 276 records (87.3%), compared to 215 records (77.9%) linked using the established CHeReL deterministic linkage approach. Both methods linked 193 records (70%) in common, and 13 records (4.7%) were not linked by either method. Notably, 22 records (8%) were linked by CHeReL but not by the proposed method, and 26 records (9.4%) were linked by the proposed method and not CHeReL.

**Table 4.**
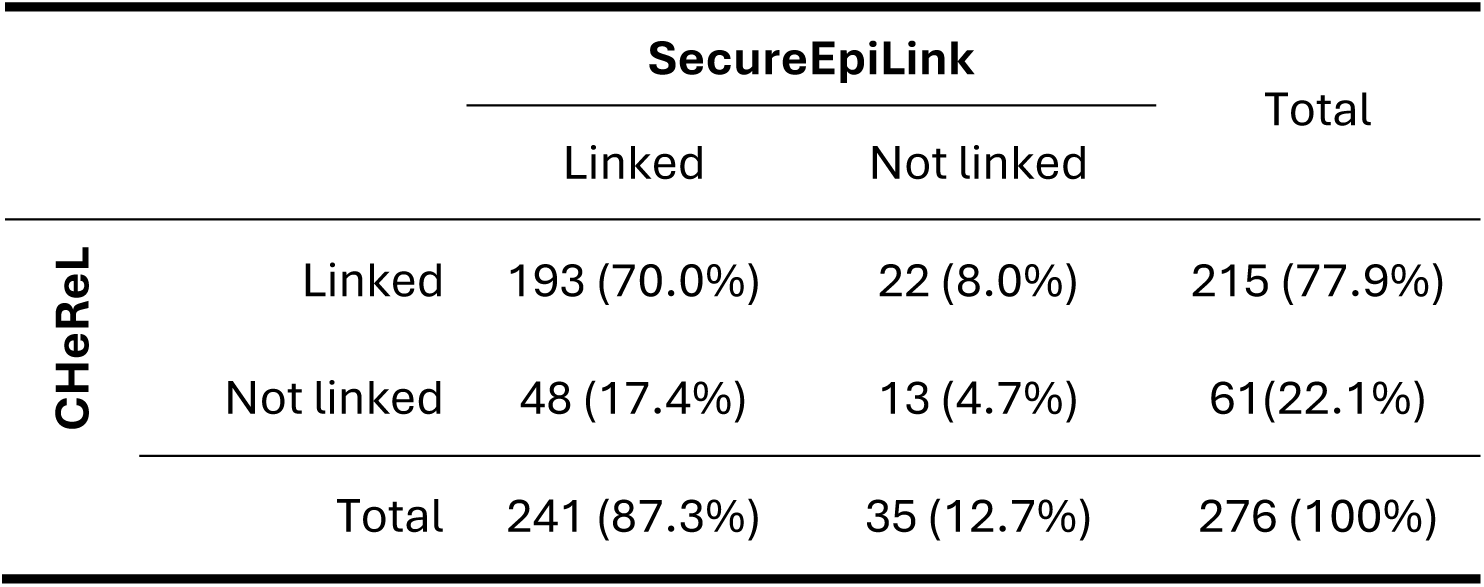
Comparison of record linkage outcomes between CHeReL and SecureEpiLink.

### Participating linkage fields analysis

An analysis of different combinations of identifiable fields as the input to an anonymised linkage identifier showed that name code and date of birth (D.O.B) fields provided the highest percentage successful linkages and lowest percentage undetermined and unsuccessful linkages for both HCV (Table 5) and HIV (Table 6).

**Table 5.**
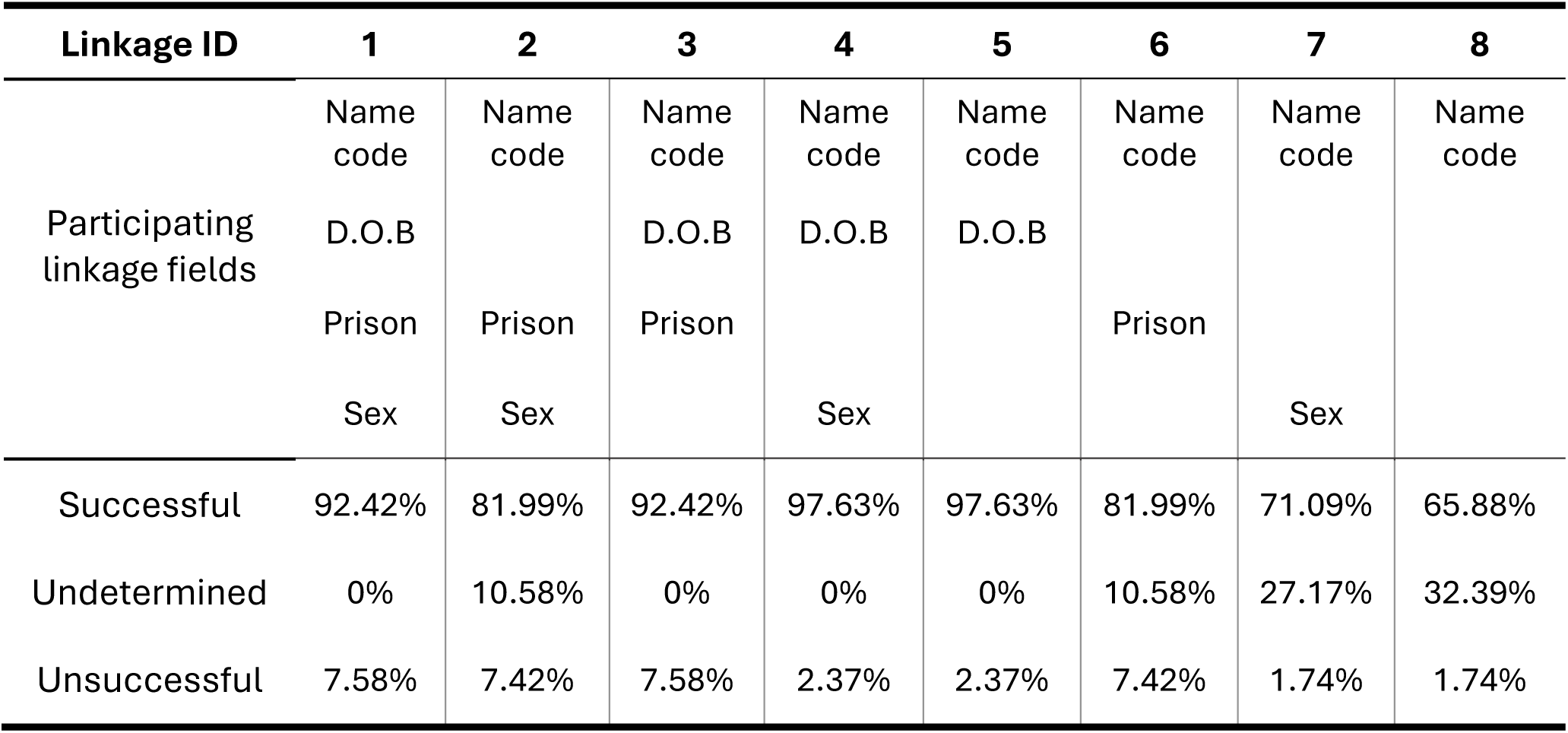
Identifiable fields inputs to the anonymised linkage identifier (ID) and corresponding SecureEpiLink linkage results for the linkage of a genomic (n=633) and notification (n=2,410) HCV dataset.

| **Linkage ID** | **1** | **2** | **3** | **4** | **5** | **6** | **7** | **8** |
|---|---|---|---|---|---|---|---|---|
| Participating linkage fields | Name code | Name code | Name code | Name code | Name code | Name code | Name code | Name code |
|  | D.O.B |  | D.O.B | D.O.B | D.O.B |  |  |  |
|  | Prison | Prison | Prison |  |  | Prison |  |  |
|  | Sex | Sex |  | Sex |  |  | Sex |  |
| Successful | 92.42% | 81.99% | 92.42% | 97.63% | 97.63% | 81.99% | 71.09% | 65.88% |
| Undetermined | 0% | 10.58% | 0% | 0% | 0% | 10.58% | 27.17% | 32.39% |
| Unsuccessful | 7.58% | 7.42% | 7.58% | 2.37% | 2.37% | 7.42% | 1.74% | 1.74% |

**Table 6.** Identifiable fields inputs to the anonymised linkage identifier (ID) and corresponding SecureEpiLink linkage results for the linkage of a genomic (n=276) and notification (n=22,187) HIV dataset.

| **Linkage ID** | **1** | **2** | **3** | **4** | **5** |
|---|---|---|---|---|---|
| Participating linkage fields | Name code<br>D.O.B<br>Postcode<br>Sex | Name code<br>Postcode<br>Sex | Name code<br>D.O.B<br>Postcode | Name code<br>D.O.B<br>Sex | Name code<br>D.O.B |
| Successful | 40.22% | 40.22% | 40.58% | 87.32% | 88.77% |
| Undetermined | 0% | 1.45% | 0% | 0% | 0% |
| Unsuccessful | 59.78% | 58.33% | 59.42% | 12.68% | 11.23% |

**Table 7.** Linkage transaction result between a single mock public health unit node and two mock laboratory nodes.

| **Mock data node** | **Public Health Unit** | **Laboratory 1** | **Laboratory 2** |
|---|---|---|---|
| Record type | Notification | Genomic | Genomic |
| Total records | 347 | 166 | 181 |
| Successful linkages | 330 (95.1%) | 156 (94%) | 174 (96.1%) |

The highest successful linkage percentages were 97.63% and 88.77%, for HCV and HIV respectively, where successful linkage is defined as the number of genomic records uniquely linked with a single notification record. For the HCV datasets, name code alone was insufficient to link records, with the percentage of successful linkage being the lowest out of all the 8 linkage identifiers generated. The addition of sex resulted in either an unchanged linkage success result or decreased the linkage success in the case of the HIV datasets. Similarly, the addition of any additional participating linkage fields resulted in higher percentages of both undetermined and unsuccessful linkage results.

Although additional linkage fields increased the uniqueness of a linkage identifier, as shown by the decrease in undetermined linkages, or linkage collisions (Tables 4 and 5). This was accompanied by an overall decrease in linkage success, which is to be expected, as a greater level of agreement is required between two datasets to achieve successful linkage.

Please note that as these are real datasets with no ground truth in relation to if a record is successfully linked, the difference between an increase in true matches and an increase in false matches cannot be distinguished. As a result of this, linkage success described here should not be directly read as an indication of linkage accuracy.

### Distributed data linkage platform

SecureEpiLink was developed to support two types of data collecting organisations, or data nodes: pathology laboratories and public health departments/units. Laboratory data nodes are designed to represent pathology laboratories responsible for pathogen sequencing, and public health unit nodes to represent data collecting organisations storing public health notification metadata. SecureEpiLink was based on a distributed architecture model in which each data collecting organisation, either laboratory or public health notification unit, would manage its own data node. Within-jurisdiction data flow between these two node types would be bi-directional and cooperative, with data linkage completed at the laboratory node, and sequence metadata and files (fasta) being shared from the laboratory node to the public health unit node upon successful linkage. The linkage, data and analysis flow between public health and laboratory nodes was designed in three key stages: data node initialisation, data linkage and sharing, and bioinformatic analysis and report generation (Figure 2).

**Figure 2.**
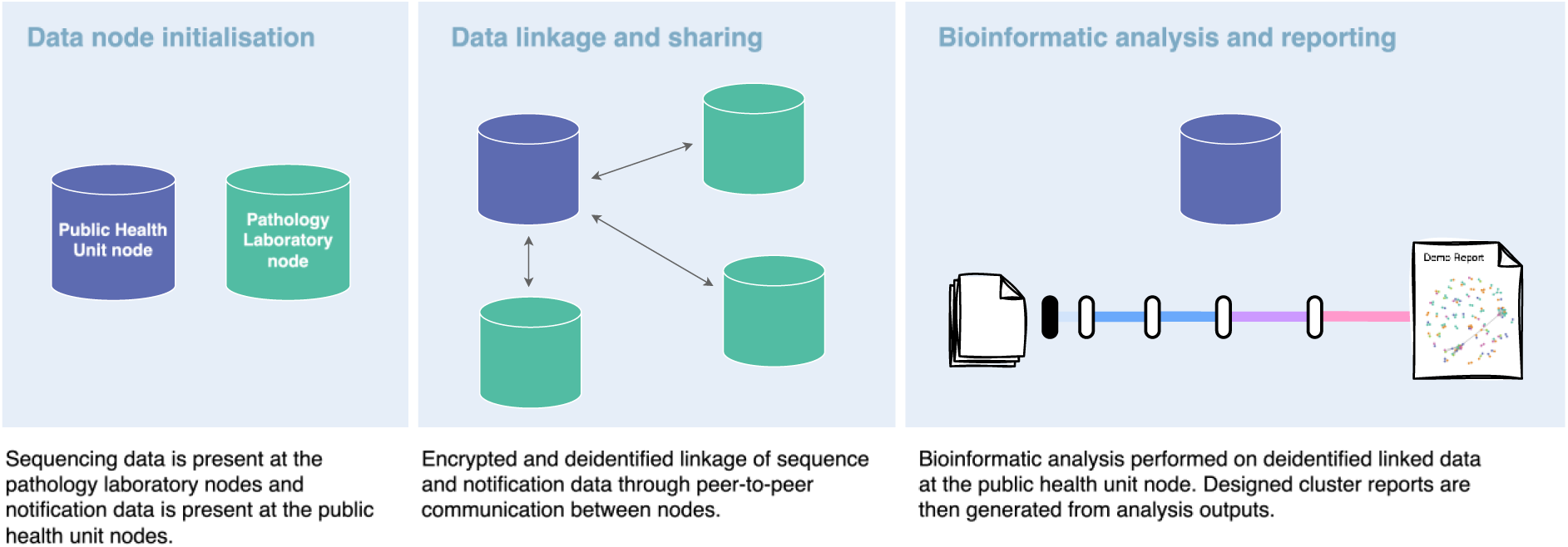
High level data flow. Designed linkage, data sharing, analysis and reporting flow between public health and laboratory nodes.

Peer-to-peer communication between data nodes to facilitate the data linkage and sharing component was managed via RESTful APIs, an industry standard for building public facing APIs and web services, where each node operates as an independent service. These APIs were secured using HTTPS, behind a reverse proxy. Between-node requests were authenticated using a unique key assigned to each registered node, intending only to demonstrate the approach in a prototype setting. Production deployments would require certificates from a trusted certificate authority, stronger authentication mechanisms, and secure key management. Data linkage and sharing via these APIs was designed as a data linkage transaction between a Laboratory node and a Public Health Unit node (Figure 3). A data linkage transaction is initiated from the laboratory node, where a one-way cryptographic hash function is used to de-identify the participant data and generate linkage identifiers at both node types. These linkage identifiers are generated from the combination of a record’s sensitive fields, name code and date of birth, and a salt, which are then hashed or transformed into non-decodable form. During linkage only the hashed, anonymised identifiers are shared between nodes, and once linkage has occurred, only the non-sensitive fields are shared between nodes. After the completion of a linkage transaction, linked records and genomic sequence files are stored at the public health unit node. The salt can then be destroyed, ensuring privacy and adding a layer of security by preventing further linkage or re-identification. The entire linkage and data sharing process is managed within one transaction which is identifiable through a correlation identifier.

**Figure 3.**
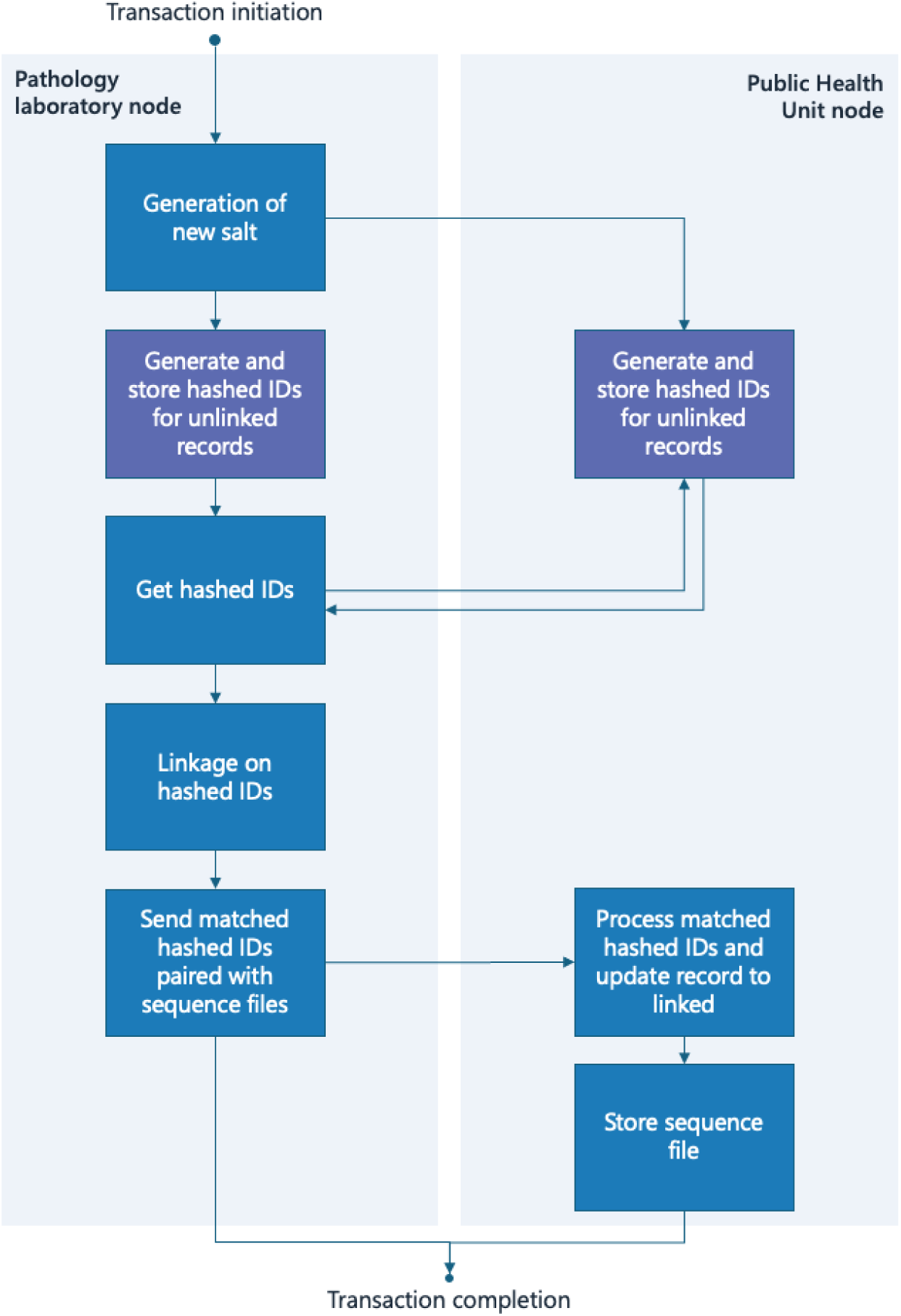
**Key stages of a data linkage and data sharing transaction**. Transaction involves peer-to-peer communication via APIs (Supplementary Figure 1) between a single laboratory node (left) and a single public health unit node (right).

The database schema was designed to support both node types, laboratory and public health unit, with each node type maintaining its own consistent data model (Supplementary Figure 2). Several common entities were shared between node types to ensure interoperability. Server-side checks were included to ensure data integrity, supported by the development of mock upload templates and data dictionaries for HIV genomic and notification records. Mock upload templates and data dictionaries allow users to prepare and format their data in the expected structure, reducing the probability of data-integrity errors reaching the database. Uploaded datasets were stored across the Records and Metadata tables, with flexible configuration of metadata, linkage and sharing fields. The Public Health Unit node data model contained two additional tables for storing the linked, anonymised records and associated metadata. Linked records in these tables were identified with new, newly generated External Identifiers, unique to SecureEpiLink.

The HIV simulation datasets, including genomic sequence files, were successfully uploaded across two mock laboratory type data nodes and one mock public health unit data node. A linkage transaction was then initiated between each laboratory data node and the public health unit node to link genomic and notification records, where 330 out of a total of 347 records (95.1%) (Table 7) across all three data nodes were successfully linked. All successfully linked records and genomic sequence files were stored at the mock public health unit node.

### Report generation pipeline

SecureEpiLink was enhanced to support the execution of an analysis pipeline, generating a pre-designed HTML report summarizing linked records and metadata stored at the public health unit node. This process is initiated by a POST API request that triggers a SnakeMake pipeline, followed by a subsequent POST API request that generates the final HTML report. Separating the pipeline and report generation steps allows for greater future implementation flexibility, particularly in scenarios where reports need to be regenerated without repeating expensive bioinformatic computations, or varying report versions need to be generated for different stakeholders based on the same bioinformatic outputs. Example visualisations generated from the pipeline and reporting component included a hypothetical molecular cluster network, network statistic summaries and proportional distribution plots of metadata variables over 6-month time intervals (Figure 4).

**Figure 4.**
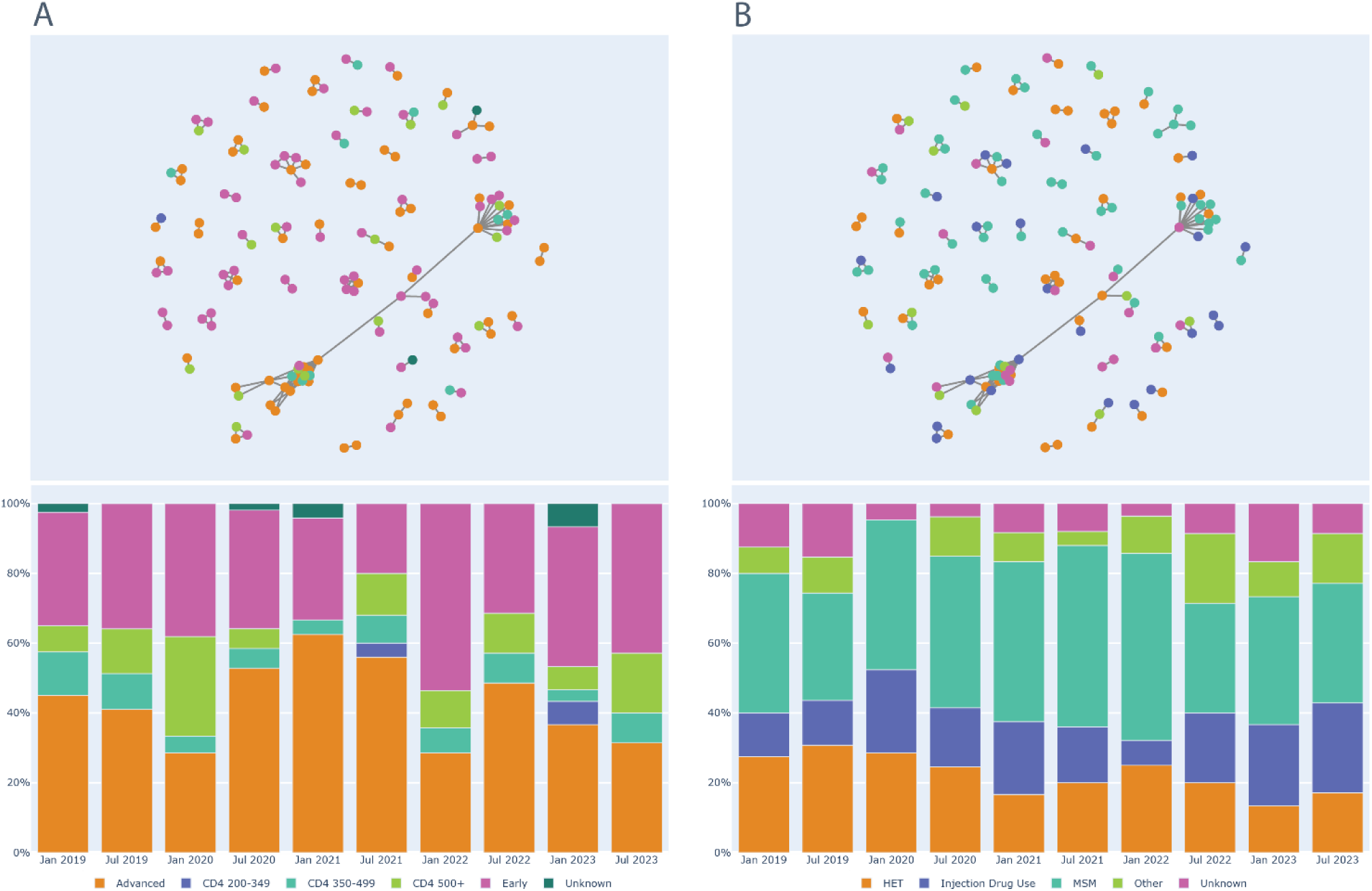
Example figures generated within the Quarto HIV report. Molecular cluster network diagram (top) and proportional distribution of overlayed metadata over time (bottom), coloured by stage of infection (A) and risk of exposure (B). HET, heterosexual; MSM, men who have sex with men. Note that data shown is simulated with a random allocation of stages of infection and risks of exposure.

Although the linkage, and the analysis and reporting components of the system were both logically integrated, sharing the same node-specific MySQL database, both components were segregated by running of each part of the system in self-contained environments (Supplementary Figure 3). This supports inter-component data consistency, while maintaining modularity and flexibility for future development and scaling.

The proof-of-concept linkage platform, with analysis and reporting components, is provided in a minimal, open-source format to allow for replication of the linkage, pipeline and reporting outputs. There are five key steps in setting up and running a minimal linkage, analysis and reporting example using the provided testing dataset:

1. Creating a mock Public Health node and two mock Pathology Laboratory nodes
2. Uploading notification records, sequence records and genomic files to each of the three nodes
3. Initiating a linkage transaction between both Pathology Laboratory nodes and the Public Health node
4. Initiating analysis pipeline at the Public Health Unit node
5. Generating an example templated report at the Public Health Unit node

The source code, detailed running instructions and documentation to test the proof-of-concept is available in a publicly accessible Git repository [21].

## Discussion

The showcased privacy-preserving linkage protocol, which enables record matching without the need for sharing of sensitive and identifiable fields, was successfully implemented within a proof-of-concept distributed data linkage and sharing platform. We demonstrated feasibility of data linkage and sharing across independently maintained datasets, in simulated conditions. This emulates the scenario where each data collection organisation manages its own data autonomously, while SecureEpiLink directly integrates linkage functionality to support timelier, near real-time data integration. Such an approach could enable faster detection of genetic clusters and more responsive public health action compared to current retrospective and centralised linkage workflows.

While distributed platforms allow for effective data storage, linkage and sharing capabilities, there is significant added complexity in development, implementation and maintenance of an operationalised distributed platform. Mechanisms need to be introduced, managed and maintained to standardise storage, queries and results across possibly heterogeneous data sources with varying policy and governance requirements. Similarly, mechanisms for code versioning, deployment of new features and critical updates, and conflict resolution are required to ensure synchronisation and consistency across all nodes within the distributed network. Secure communication protocols also need to be maintained not only within data nodes, like in a centralised system, but also between distributed nodes. The key considerations, impacts and approaches are summarised in Table 8. Despite these operational and technical challenges, our proof-of-concept implementation demonstrates that distributed linkage using hashed anonymised identifiers, specifically name code and date of birth, can achieve success rates comparable to established protocols.

**Table 8.** Key implementation considerations for distributed systems such as SecureEpiLink.

| **Consideration** | **Impact** | **Approach** |
|---|---|---|
| Data standardisation and heterogeneity | Nodes may store data from different source and in different formats, yet data model, queries and bioinformatic results must be consistent across the distributed system for successful linkage and accurate public health action. | Allow for flexible per-node store of data, while agreeing and validating on common data formats and query standards across all nodes within the distributed system. |
| Policy and governance | Data storage, sharing and privacy policies can vary heavily across organisations, making it challenging to maintain one solution that suits all.<br>Concerns around data ownership relating to intellectual property and research opportunities.<br>Control over public health messaging regarding outbreaks. | Establish agreements between data collecting organisations to define data storage and retention, linkage and sharing policies.<br>Establish agreements regarding use of data shared between organisations.<br>Link genomic and epidemiological data at regular intervals within public health units reporting to health departments in each jurisdiction. |
| Platform operationalisation and maintenance | Operating and maintaining a distributed system requires consistent deployments, updates and monitoring across many nodes without the use of a central authority. | Build dependable automation pipelines, strong version control protocols, and ensure flexibility to scale is at the forefront of design. |
| Security and authentication | As a distributed network grows, so does the importance of security, as each additional node increases the number of potential points of compromise. | Use techniques such as certificated-based authentication methods, central node registries, role-based access control, encrypted communication, and detailed system logging to strengthen the platform. |

The ability to perform de-identified linkage is particularly relevant for pathogens such as HIV, which present significant ethical and legal considerations associated with stigma and criminalisation [13, 14]. These factors require strict separation of identifiable information from analytical datasets and highlight the importance of privacy-preserving methods such as the one demonstrated here.

While this approach offers clear advantages for data-security, the effectiveness of the method was found to be highly dependent on the uniqueness of the linkage field combinations, reflecting the inherently deterministic nature of hashing. In practice, our evaluation showed higher success rates on small datasets, and highlighted significant constraints with larger datasets. With the number of historical cases of newly acquired HIV reported over the last 10 years in NSW being less than 400 per year [20], the occurrence of duplicate linkage keys, and therefore failed linkages, would be negligible. As the testing datasets scaled to 12 million records, approximately 8% of records had duplicate identifiers (Table 3), highlighting possible limitations in scalability to national or international implementations. This observation is consistent with the birthday problem, which describes the quadratic growth of the absolute count of accidental collisions, in this case different records sharing the same name code and date of birth, as the dataset grows by a factor of 10. This limitation in using simply name code and date of birth for linking records reliably in large datasets could be mitigated by the inclusion of more linkage fields, or more complete information in the fields.

A further constraint of hash-based deterministic linkage is its susceptibility to minor data entry errors and character-level differences, such as accents, apostrophes and hyphens within name fields. Fields also vary in how prone they are to mismatch. Postcodes are particularly prone to mismatch for two reasons: they are time-varying variables that may change between the date of testing and the date of notification, or they can often be unreliable with postcodes of medical clinics being recorded rather than an individual’s residential postcodes. Prison, or incarceration facility, names are also prone to mismatch. They are often recorded as free text fields, with significant variation in spelling and formatting, and are at risk of changing over time due to nomenclature shifts, facility mergers and administrative rebranding. Identifiers used in linkage should therefore be uniform, with rigorous data collection and standardisation practices and data checking implemented at the point of data entry.

A key limitation of this study is that the linkage of HIV notification records was done without our direct access to the NCIMS database, and CHeReL’s linkage process is closed source, resulting in an inability to access the missing value counts or note the characteristics of unlinked records. Therefore, the extent of the effect of missing values on HIV linkage success, and understanding the factors driving failed linkages, is unclear. In the linkage of HCV data, missing and inconsistent linkage fields in records were a common cause of unlinked records, and while this is likely a cause of unlinked records for the HIV data, it cannot be determined definitively.

While optimising linkage outcomes was not the primary aim of this study, linkage success remains a critical factor of the comprehensiveness and quality of the final linked datasets, which in turn directly influences downstream genomic and epidemiological analyses to guide public health interventions. The adoption of data guidelines will be essential to enable a robust and reproducible implementation of this linkage protocol and distributed data linkage and sharing platform. This includes community-agreed metadata standards, controlled vocabularies, and shared ontologies which support interoperability and reusability while still allowing for sensitive data to be tightly access-controlled.

Addressing the key constraints in relation to both the linkage protocol and platform operationalisation will be essential to enabling successful multi-jurisdictional implementation of a distributed linkage, analysis and reporting platform such as SecureEpiLink. Future work should focus on several key areas including the exploration of alternate linkage approaches that address scalability limitations, aligning with policy and public health specialists, and operationalising SecureEpiLink with robust orchestration tools.

Furthermore, SecureEpiLink should be extended to incorporate critical features and functionality that ensure usability, such as administrative interfaces for managing key aspects such as pathogen record data dictionaries and data upload templates. Capturing these requirements ensures that the system can be reliably used, maintained, and governed in real-world settings, while supporting reproducibility and accountability across analyses.

## Conclusion

Distributed, privacy-preserving data sharing and linkage showcases a promising alternative for traditional centralised systems, allowing for data collecting organisations to manage their own data, while also providing a strong foundation to genomic and epidemiological analysis and reporting on linked datasets. Scaling SecureEpiLink across jurisdictions will require the integration of SecureEpiLink with existing health data systems, development of secure methods for verifying user identity and protecting data, and structured processes for system roll out, maintenance, and updates.

## Data Availability

Simulated data is available from https://github.com/jolenefarrell/SecureEpiLink/tree/master/automation/test_data

## Data Availability

All data produced in the present study are available upon reasonable request to the authors

https://github.com/jolenefarrell/SecureEpiLink

## Acknowledgements

This research was supported by The University of Melbourne’s Research Computing Services and the Petascale Campus Initiative. We would like to acknowledge the AusTrakka program for its support.

## H2Seq Data Management Working Group

Lachlan Coin^1^, Ansari Shaik^2^, Claire Vajdic^2^, Jonathan King^2^, Chaturaka Rodrigo^5^, Alison Castley^7^, Cipriano Martinez^8^, Daneeta Hennessy^9^, David Whiley^10^, Deborah Williamson^11^, Chuan Lim^11^, Leon Caly^11^, Ammar Aziz^11^, George Taiaroa^11^, Jodie D’Costa^12^, Doris Chibo^12^, Karen Hawke^13^, Leon Patrick McNally^14^, Steven Nigro^15^, Rebecca Stephenson^15^, Shannon Melody^16^, Sebastiaan Van Hal^17^, Simon Reid^18^, Torsten Seemann^19^, Tuyet Hoang^19^, Lex Leong^20^

7 West Australia Department of Health

8 National Association of People Living with HIV Australia

9 Victorian Department of Health

10 University of Queensland Centre for Clinical Research

11 Department of Infectious Diseases, University of Melbourne

12 Victorian Infectious Disease Reference Laboratory

13 South Australian Health and Medical Research Institute

14 St Vincents Health Australia

15 New South Wales Department of Health

16 Tasmanian Department of Health

17 Royal Princes Alfred Hospital, Sydney

18 University of Queensland

19 Centre for Pathogen Genomics, University of Melbourne

20 SA Pathology

