## Supplementary tables and Figures for "A distributed, privacy-preserving platform for linkage of epidemiological data with pathogen genome sequences"

**Supplementary Table 1**. Key thresholds and parameters, including corresponding descriptions or justifications, utilised in simple SnakeMake pipeline running HIV-TRACE.

| **HIV-TRACE parameters** | | |
|---|---|---|
| Parameter | Value | Description/justification |
| -r (reference) | HXB2_prrt | HXB2 is the standard reference genome in the literature. |
| -a (ambiguities) | skip | Expectation that no sequences assembled with ambiguous bases to be submitted to the platform. Skip any that may be present to reduce sources of noise. |
| -t (distance threshold) | 0.03 | Appropriate threshold to utilise for more divergent clusters as present historically in NSW. |
| -s (strip resistant sites) | lewis | Use 'lewis' dataset to filter our drug resistant sites for more accurate distance calculation. |
| -m (min overlap) | 500 | - |

**Supplementary Table 2**. Generated sequence metadata fields for the HIV simulation genomic dataset.

| Field | Description | Valid values (Sampling probability) |
|---|---|---|
| Internal ID | Unique string identifier for the record | - |
| Name code | First two characters of the individual’s first name and last name | - |
| Date of birth | Individual’s recorded date of birth (DD/MM/YYYY) | 01/01/1950 - 01/01/2010 |
| Postcode | Metropolitan and regional NSW postcodes | 2000-2300 (0.95)<br>2301-2599 (0.5) |
| Sequence ID | Identifier assigned to the sequence at the data collecting organisation | - |
| Date of collection | Date of specimen collection | 01/01/2019 - 01/01/2024 |

**Supplementary Table 3**. Generated notification metadata fields for the HIV simulation notification dataset.

| **Field** | **Description** | **Valid values (Sampling probability)** |
|---|---|---|
| Internal ID | Unique string identifier for the record | - |
| Name code | First two characters of the individual's first name and last name | - |
| Date of birth | Individual's recorded date of birth | 01/01/1950 - 01/01/2010 |
| Postcode | Metropolitan and regional NSW postcodes | 2000-2300 (0.95)<br>2301-2599 (0.5) |
| Date of notification | Date of initial diagnosis notification to the relevant health department | 01/01/2019 - 01/01/2024 |
| Age at diagnosis | Calculated from the date of notification and date of birth fields | 0-9, 10-19, 20-29, 30-39,<br>40-49, 50-59, 60+ |
| Stage of infection | Stage of infection at time of notification | Early (0.3)<br>CD4 500+ (0.1)<br>CD4 350-499 (0.08)<br>CD4 200-349 (0.01)<br>Advanced (0.4)<br>Unknown (0.01) |
| Subtype | HIV-1 subtypes | B (0.5)<br>CRF01_AE (0.2)<br>C (0.1)<br>Recombinant (0.1)<br>Other (0.1) |

| | | |
|---|---|---|
| Risk of exposure | Reported HIV risk exposure | MSM (0.4)<br>HET (0.2)<br>Injection Drug Use (0.2)<br>Other (0.1)<br>Unknown (0.1) |
| Testing history | Occurrence of a negative or indeterminate western blot result | Neg. or ind. ≤12 months (0.35)<br>Neg. or ind. >12 months (0.45)<br>Never tested before (0.1)<br>Unknown (0.1) |
| Place of birth | Individual's recorded place of birth | Australia (0.4)<br>Overseas (0.6) |
| Acquisition | Likely place of HIV acquisition | Australia (0.8)<br>Overseas (0.17)<br>Unknown (0.03) |

**Supplementary Table 4.** Set of sensitive linkage fields utilised to assess data linkage success.

| HIV | | HCV | |
|---|---|---|---|
| **Column** | **Definition** | **Column** | **Definition** |
| Name code | First two characters of a first name and last name | Name code | First two characters of a first name and last name |
| Date of birth | Individual's recorded date of birth | Date of birth | Individual's recorded date of birth |
| Sex | Single character representing sex | Sex | Single character representing sex |
| Postcode | Postcode of residency | Prison | Incarceration facility at time of data collection |
| Sexual health clinic code | Code of the sexual health clinic individual was treated at | | |

**Supplementary Table 5**. Identifiable fields inputs to the anonymised linkage identifier (linkage ID) for HIV and HCV.

| HCV | | | | | | | | |
|---|---|---|---|---|---|---|---|---|
| Linkage ID | 1 | 2 | 3 | 4 | 5 | 6 | 7 | 8 |
| Linkage fields | Name code<br>D.O.B<br>Prison<br>Sex | Name code<br>Prison<br>Sex | Name code<br>D.O.B<br>Prison | Name code<br>D.O.B<br>Sex | Name code<br>D.O.B | Name code<br>Prison | Name code<br>Sex | Name code |

| HIV | | | | | |
|---|---|---|---|---|---|
| Linkage ID | 1 | 2 | 3 | 4 | 5 |
| Linkage fields | Name code<br>D.O.B<br>Postcode<br>Sex | Name code<br>Postcode<br>Sex | Name code<br>D.O.B<br>Postcode | Name code<br>D.O.B<br>Sex | Name code<br>D.O.B |

**Supplementary Table 6.** Summary of each condition tested, where both dataset size and name code diversity are varied. Ethnic category is defined as either diverse or non-diverse.

| Condition | Number of records | Ethnic category | Pseudo-data entry errors | Date sampling range |
|---|---|---|---|---|
| 1 | 12,000 | Non-diverse | 16.67% | 01/01/1950 - 01/01/2010 |
| 2 | | Diverse | | |
| 3 | 120,000 | Non-diverse | | |
| 4 | | Diverse | | |
| 5 | 1,200,000 | Non-diverse | | |
| 6 | | Diverse | | |
| 7 | 12,000,000 | Non-diverse | | |
| 8 | | Diverse | | |

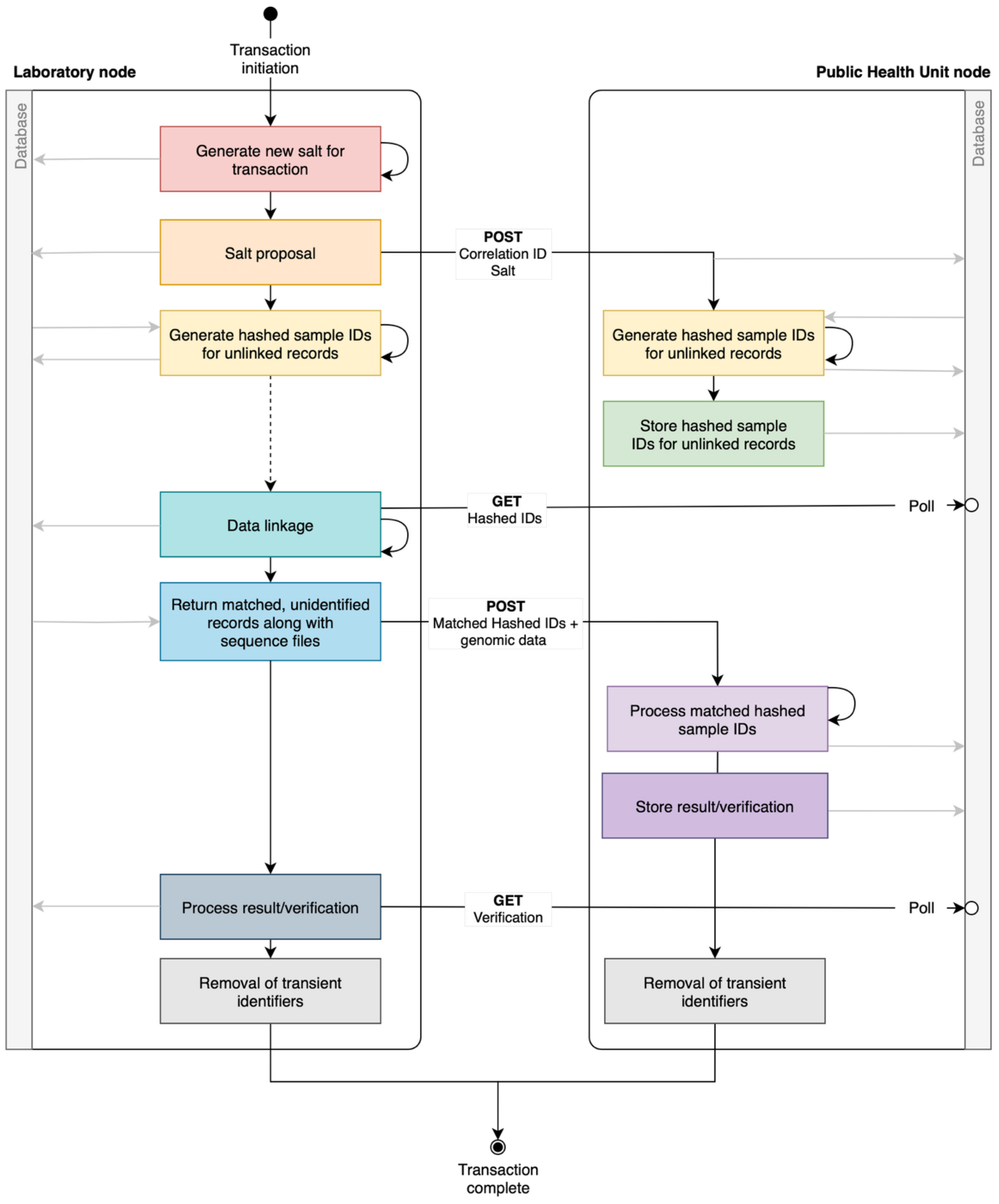


**Supplementary Figure 1.** Detailed key stages of a data linkage and sharing transaction between a laboratory node (left) and public health unit node (right), including key API requests involved.

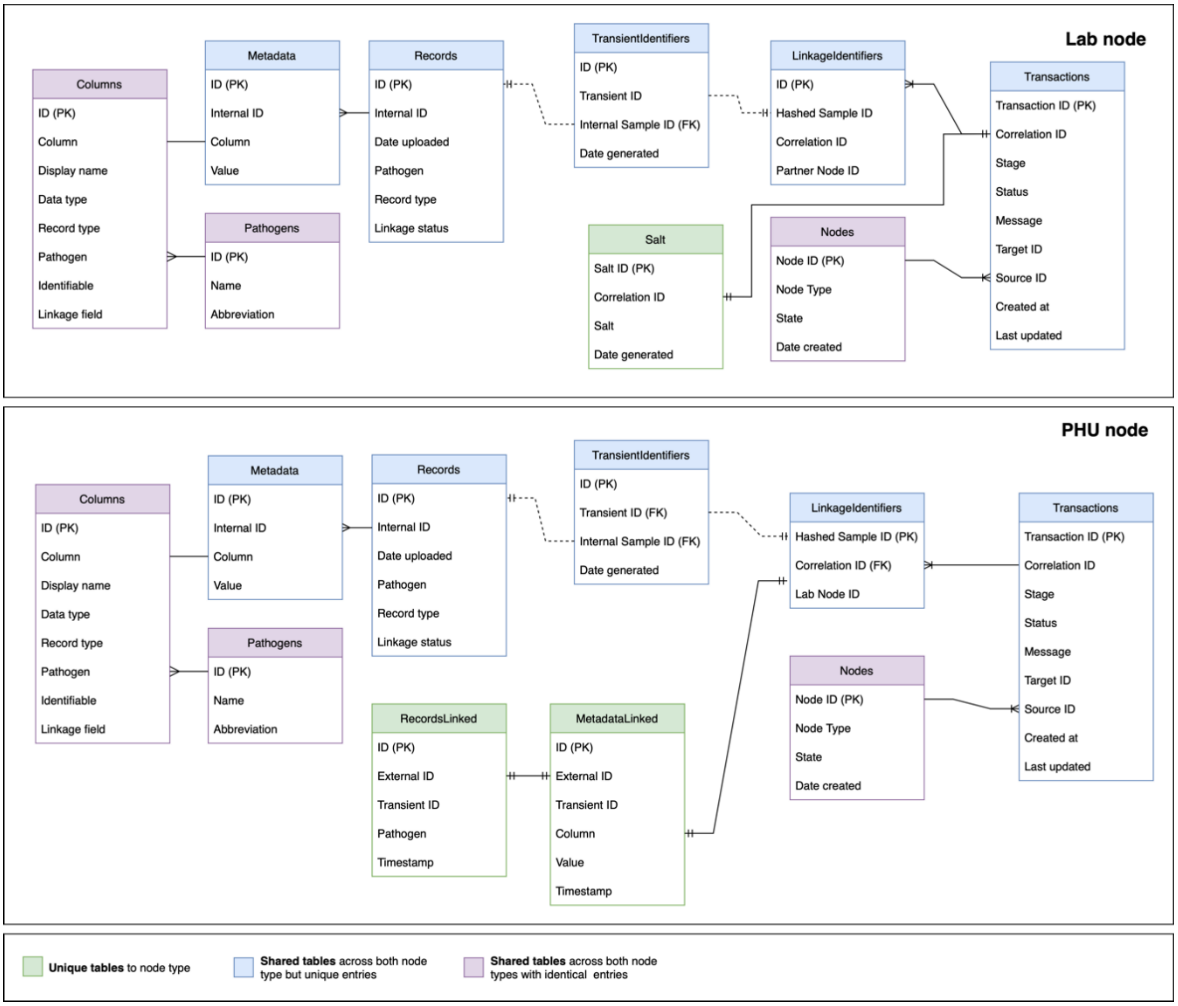


**Supplementary Figure 2.** Entity Relationship Diagram representing the data model for both the Laboratory node (top) and Public Health Unit node (bottom).

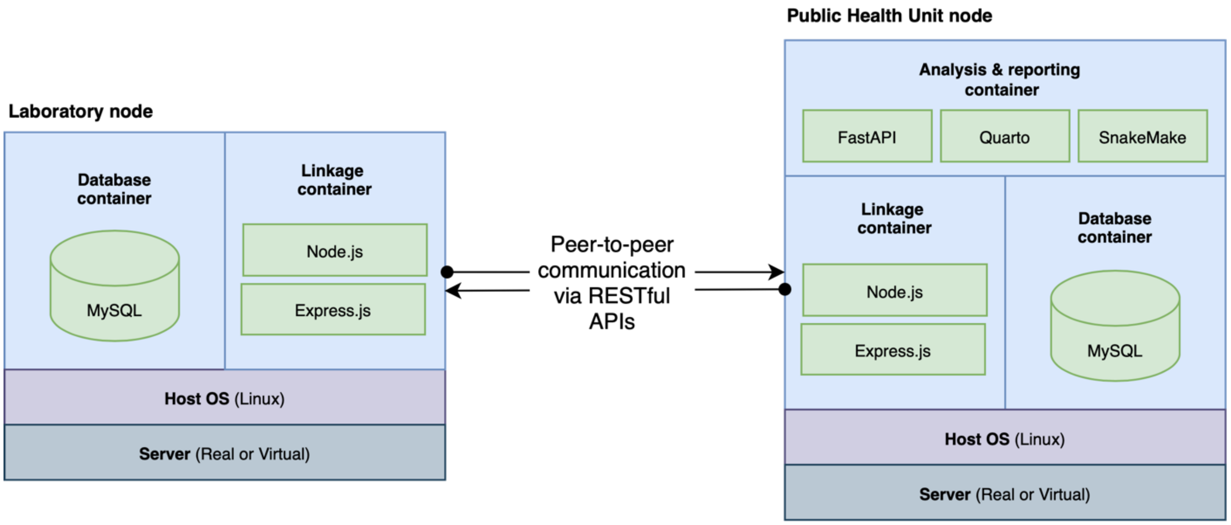


**Supplementary Figure 3**. High-level architecture diagram of both a laboratory node (left) and a public health unit node (right).
